# Connectivity between the central executive and salience networks normalizes with exposure-focused CBT in pediatric anxiety

**DOI:** 10.64898/2026.01.28.26345061

**Authors:** Dana E. Díaz, Hannah C. Becker, Felicia A. Hardi, Adriene M. Beltz, Emily L. Bilek, Stefanie Russman Block, K. Luan Phan, Christopher S. Monk, Kate D. Fitzgerald

## Abstract

Exposure is considered the most active element of cognitive behavioral therapy (CBT) for pediatric anxiety, and its efficacy is theorized to depend on cognitive control and its supporting neural substrates (e.g., central executive [CEN], salience [SN], and default mode networks [DMN]). However, little work has identified how CBT, or exposure specifically, modulates intrinsic connectivity of these networks. Progress may be limited by heterogeneity in network connectivity in anxiety, which may obscure treatment-related effects in group-averaged analyses. This randomized clinical trial (RCT) leverages person-specific network modeling to test how exposure-focused CBT (EF-CBT) influences resting-state connectivity of cognitive control networks in pediatric anxiety, relative to an active control (relaxation mentorship training; RMT). Youth aged 7-18 years with an anxiety disorder (N = 104) or low/no anxiety (L/NA; N = 37) completed resting-state fMRI scans before being randomized to EF-CBT or RMT. Resting-state connectivity was reassessed following treatment (or commensurate time L/NA youth) in 113 participants. Changes in within-CEN, CEN-SN, and CEN-DMN density were examined using Group Iterative Multiple Model Estimation, which yields sparse, person-specific networks capturing both shared and individual connectivity structure. At baseline, anxious youth exhibited lower density within-CEN, between CEN-SN, and between CEN-DMN than L/NA youth. Treatment effects differed by intervention: EF-CBT selectively increased (i.e., normalized) CEN–SN density, whereas RMT increased within-CEN density. These findings demonstrate dissociable effects of exposure and relaxation on cognitive control network organization in pediatric anxiety. Exposure-focused CBT uniquely strengthens coordination between control and salience systems, consistent with a mechanism supporting top-down control of threat-related signals during exposure. Network-based measures of cognitive control may help identify mechanistic targets for optimizing and personalizing treatment.

**Clinical Trial Number:** NCT02810171.

## Introduction

Cognitive behavioral therapy (CBT) is the gold standard treatment for pediatric anxiety disorders, and is effective for ∼60% of those treated (Walkup et al., 2008). However, CBT can be challenging for many patients because its most active component, exposure, requires patients to repeatedly confront the very fears that cause them distress (Bilek et al., 2021; Peris et al., 2015; Whiteside et al., 2020). Successful completion of exposure tasks predicts better treatment response (Benito et al., 2018; Peris et al., 2017; Simpson et al., 2011) and depends on cognitive control, the flexible adaptation of thought and behavior. However, the extent to which cognitive control networks contribute to exposure-based CBT in pediatric anxiety remains unclear, in part due to substantial inter-individual variability of network function in psychiatric disorders (Segal et al., 2023). To address this gap, this randomized clinical trial (RCT) of anxious youth examined resting-state connectivity of cognitive control networks before and after exposure-focused CBT (EF-CBT), relative to an active comparison therapy of relaxation mentorship training (RMT).

We leverage Group Iterative Multiple Model Estimation (GIMME), a network mapping approach optimized to detect heterogeneity within functional brain networks implicated in psychopathology. In doing so, this study provides the first evidence of treatment-specific changes in connectivity of cognitive control networks in an RCT in youth with anxiety disorders.

### Neural substrates for cognitive control in youth with anxiety

Altered functional connectivity of the tripartite network system has been linked to deficits of cognitive control and is thought to contribute to the neural mechanisms underlying anxiety disorders (Fitzgerald et al., 2021; Sylvester et al., 2012). This system is characterized by three topologically and functionally distinct, but interacting groups of brain regions: the central executive (CEN), salience (SN) and default mode networks (DMN) (Menon, 2011). The CEN encompasses frontoparietal regions such as the dorsolateral prefrontal cortex and posterior parietal cortex, and is functionally connected to the SN at rest and during cognitive tasks (Cai et al., 2021; Menon & D’Esposito, 2022; Sridharan et al., 2008). The SN, which includes the dorsal anterior cingulate cortex and anterior insula, is involved in switching between CEN-based cognitive control and DMN-mediated self-focused, affective thought (Sridharan et al., 2008). By contrast, the DMN is suppressed during cognitive tasks and is anti-correlated with the CEN and SN at rest (Menon & D’Esposito, 2022).

Theoretical models and emerging data suggest that altered resting-state connectivity within the CEN, and between the CEN, SN, and DMN may contribute cognitive control impairments and related difficulty controlling repetitive negative thoughts (e.g., worry) and avoidance behaviors in anxiety disorders (Fitzgerald et al., 2021; Menon & D’Esposito, 2022; Sylvester et al., 2012). Consistent with this, anxiety has been associated with altered functional coupling between overlapping executive control networks, affective/salience networks, and the DMN (Xu et al., 2019), as well as reduced white matter integrity of fronto-limbic and fronto-parietal tracts that connect these networks (Glenn et al., 2022; Tromp et al., 2012; Wang et al., 2016). In contrast, a recent meta-analysis limited to clinically anxious populations reported no reproducible network-level alterations in resting-state connectivity, attributing null findings to substantial clinical and methodological heterogeneity across studies (Zugman et al., 2023). These mixed results suggest that anxiety-related alterations in intrinsic connectivity may not be well captured by predominant methods like group-averaged analyses, motivating approaches that can model both shared and individual-level variation.

### Brain-based mechanisms of CBT in youth with anxiety

Exposure is thought to engages cognitive control processes that enable patients to suppress reflexive avoidance in the presence of feared stimuli (Craske, 2015; Mason et al., 2016; Picó-Pérez et al., 2022). Repeated exposure practice may, in turn, improve these control processes over the course of CBT (Hadwin & Richards, 2016; Hauner et al., 2012). Emerging evidence suggests that the function and connectivity of networks supporting cognitive control may be relevant for exposure-based treatment response. For instance, in youth and adults with obsessive compulsive disorder (OCD), greater resting-state connectivity between the CEN and SN predicts better response to exposure therapy (Russman Block et al., 2022). During cognitive tasks, CBT is associated with increased CEN connectivity in OCD, and increased CEN activity in major depressive and post-traumatic stress disorders, supporting the notion that CBT may strengthen the neural capacity for cognitive control (Becker et al., 2024; Yang et al., 2018). However, evidence in pediatric anxiety is mixed. During emotion-laden cognitive or attentional tasks, anxious youth show increased pre-to-post-CBT engagement in regions that interact with the CEN to support affective and regulatory control, including the rostral anterior cingulate cortex (Burkhouse et al., 2018) and ventrolateral prefrontal cortex (Maslowsky et al., 2010). Conversely, pre-to-post-CBT decreases in CEN activity have been reported during a similar threat attention task (Haller et al., 2024). However, to our knowledge, no RCT studies have identified CBT-related changes in resting-state connectivity of cognitive control networks in pediatric anxiety.

### Heterogeneity in neural substrates of pediatric anxiety and CBT response

One likely explanation for the absence of consistent CBT-related changes in resting-state connectivity is the substantial clinical and neural heterogeneity observed in pediatric anxiety. Youth anxiety is marked by high rates of comorbidity, dynamic symptom expression, and shifting symptom profiles across development (e.g., separation anxiety in childhood predicting panic disorder in adolescence), resulting in significant cross-sectional heterogeneity (Kendall et al., 2010; Pine et al. 1996; Nelemans et al., 2014; Rapee et al., 2013; Speranza et al. 2023). Consistent with this clinical variability, studies have identified distinct neurobiological profiles within youth internalizing disorders and adult anxiety that associate with differences in cognitive function (Ge et al. 2023; Kaczkurkin et al., 2020; Price et al., 2020). Moreover, systematic reviews CBT in pediatric anxiety highlight substantial variability in prefrontal and network-level changes in brain activation and connectivity, suggesting that averaging across individuals may obscure meaningful treatment-related neural effects (Aupperle et al., 2024).

Accordingly, the present study leverages GIMME (Gates & Molenaar, 2012) to examine resting-state functional connectivity of the CEN with other tripartite networks (CEN, SN, DMN) in anxious youth before and after exposure-based CBT. Conventional resting-state approaches quantify the strength of pairwise correlations between seeds or regions and often rely on group-averaging or arbitrary thresholding, which can mask meaningful individual heterogeneity (Molenaar, 2004; Poldrack, 2017). By contrast, GIMME creates sparse, person-specific directed networks by estimating only the connections that are required to explain an individual’s brain dynamics, while also estimating connections that are meaningful most participants’ networks. After estimating all person-specific GIMME connections, networks can be characterized via applications of graph theoretical metrics (see Beltz & Gates, 2017). Here, we use network density -- the proportion of model-estimated connections expressed within or between networks (over the number of all estimated connections in each individual’s sparse network). Density indexes the degree of network integration rather than the magnitude of individual correlation, enabling us to capture individual differences network organization without assuming uniform connections or patterns across youth.

### Current study

This RCT tests the effects of exposure-focused CBT (EF-CBT) compared to a relaxation-based comparison (relaxation mentorship training; RMT) on resting-state connectivity of cognitive control networks in pediatric anxiety. Although relaxation is typically delivered with CBT (Walkup et al., 2008), it is less effective than exposure (Bilek et al., 2021, 2025; Peris et al., 2015; Walkup et al., 2008). Using an active comparison intervention enabled us to control for non-specific treatment effects (e.g., time, practice, therapist contact) and isolate effects specific to EF-CBT. Functional connectivity was estimated at pre- and post-treatment with confirmatory Subgrouping-GIMME (CS-GIMME; Beltz & Gates, 2017; Gates et al., 2017), which estimates person-specific networks prioritizing not only group-level connections, but also connections shared among *a priori* defined participant subgroups, before estimating individual-level connections. At baseline, we hypothesized that anxious youth would display weaker between CEN-SN and within-CEN density, and greater (i.e., less segregated) between CEN-DMN density than youth with low or no anxiety (L/NA). Building from prior work suggesting that exposure engages cognitive control and its neural substrates (Burkhouse et al., 2018; Fitzgerald et al., 2021; Mason et al., 2016; Yang et al., 2018), we hypothesized that EF-CBT would be associated with greater increases in between CEN-SN and within-CEN density, and greater decreases in CEN-DMN density, relative to RMT.

## Methods

### Participants

The clinical trial sample included 139 pediatric patients (ages 7-18) with one or more anxiety disorders (e.g., social, generalized, separation anxiety) and 44 participants with L/NA (*Figure S1*). All participants received a clinician-administered interview with the DSM-5 version of the Kiddie Schedule for Affective Disorders and Schizophrenia–Present and Lifetime Version (KSADS-PL) to establish: 1) presence of an anxiety disorder in patients and 2) absence of any DSM-5 current or lifetime disorder in L/NA participants. In patients, some comorbid disorders (e.g., obsessive compulsive, attention-deficit/hyperactivity [ADHD]) were allowed, if anxiety was the primary source of interference and distress. Patients were excluded for: current diagnoses of major depression or substance/alcohol abuse/dependence; lifetime diagnoses of bipolar and psychotic disorders, intellectual disability, autism spectrum disorder, and post-traumatic stress disorder; acute suicidal risk; or if currently receiving psychotherapy or psychotropic medication (except medication for ADHD [n = 2], which was withheld during scanning). Standard inclusion/exclusion criteria for neuroimaging studies were also applied (see Supporting Information). Participants and guardians provided informed assent/written consent to participate, in compliance with the Michigan Medicine Institutional Review Board (HUM00118950).

### Trial registration

The trial, which follows the CONSORT Checklist (Figure S1), is titled *Dimensional Brain Behavior Predictors of CBT Outcomes in Pediatric Anxiety* and can be found at https://clinicaltrials.gov/study/NCT02810171. The study was first posted to the Clinical Trials Registry in July 2016 and is associated with the following trial registration number: NCT02810171.

### Patient randomization and treatment

Patients were randomized in a 2:1 ratio, stratified by sex and age, to receive up to 12 sessions of EF-CBT or RMT, delivered over a maximum of 16 weeks. EF-CBT prioritized exposure (sessions 4-12) and included initial psychoeducation and a single session devoted to cognitive restructuring. RMT taught relaxation strategies (e.g., structured diaphragmatic breathing and muscle relaxation), and omitted exposure and cognitive restructuring (Bilek et al., 2021). Scans were collected ∼16 weeks apart for both patients and L/NA youth.

Change in anxiety symptoms was calculated as the difference between baseline and post-treatment scores on the clinician-administered Pediatric Anxiety Rating Scale (PARS), which was evaluated by independent raters blinded to treatment condition. Global symptom improvement was also assessed by clinicians using the Clinical Global Impression of Improvement scale (CGI-I), and participants provided self-report on the Children’s Depression Inventory (see Supporting Information). The PARS was the primary measure of symptom severity and the CGI-I was assessed for comparison with prior clinical trials (Bilek et al., 2021; Peris et al., 2015).

### fMRI processing

Two five-minute resting-state fMRI scans were collected per standard protocol, following acquisition of low- and high-resolution anatomical sequences for co-registration and normalization of resting-state data. Participants with excess motion (>0.5mm mean framewise displacement; FD) were excluded from analyses before preprocessing (*Figure S1*). Resting-state timeseries were pre-processed using standard pipelines, constructed in FSL (v6.0), as in prior work (Beltz et al., 2019; see Supporting Information). Next, timeseries data from both runs of the remaining, usable scans were concatenated and extracted from 14 regions-of-interest (ROIs; *Table S2*), selected for membership in large-scale functional networks: the CEN, SN (including amygdala), and DMN. Each ROI was a 4mm-radius sphere, centered around Montreal Neurological Institute (MNI) coordinates extracted from NeuroSynth (Yarkoni et al., 2011), a meta-analytic tool that establishes peak activation for brain ROIs (pre-registered: https://osf.io/6xef8/).

### CS-GIMME

Confirmatory subgrouping GIMME (CS-GIMME; Gates et al., 2017) was applied to estimate directed functional connectivity among ROIs at baseline and follow-up for all participants. CS-GIMME estimates both contemporaneous and first-order lagged connections within a unified structural equation modeling framework, allowing characterization of instantaneous (i.e., within a TR) and time-delayed (i.e., from one TR to the next) dependencies in brain activity. This method overcomes core limitations of standard approaches that average contemporaneous connections across participants and mask meaningful heterogeneity, or that analyze each participant separately, preventing the identification of shared network organization. By contrast, CS-GIMME accounts for group-level (and in this case, subgroup-level) similarity and individual heterogeneity, often outperforming other functioning connectivity approaches, especially when data are heterogeneous (Gates & Molenaar, 2012; Lane et al., 2019; Mumford & Ramsey, 2014). The confirmatory-subgrouping feature was used to align model specification to the RCT design, allowing us to evaluate subgroup-specific change from baseline to follow-up, while retaining connections that were shared across the majority of the sample. The inclusion of both clinically affected and L/NA youth promotes the estimation of network connections that generalize beyond diagnostic boundaries.

Model-building occurs in a stepwise process. Critically, CS-GIMME does not estimate a fully connected node-by-node network for each participant. Instead, it begins with a null model that includes only autoregressive effects (each ROI predicting itself from the previous TR). The algorithm then identifies whether there is a candidate connection (i.e., one node predicting another at the same or next TR) that significantly improves model fit using Lagrange multiplier tests (which are chi-square distributed), employing multiple comparison corrections.

Connections that are significant and that improve fit for at least 75% of participants are iteratively estimated within the unique networks of all participants as group-level connections. Next, the same procedure is applied separately within each predefined subgroup (EF-CBT, RMT, L/NA). Once group- and subgroup-level structure is set, additional connections are estimated within each participant’s network until their model fits well, according to at least two of four predefined indices: comparative fit index ≥ 0.95; non-normed fit index ≥ 0.95; root mean squared error of approximation ≤ 0.05; standardized root mean residual ≤ 0.05 (Gates et al., 2014; Gates & Molenaar, 2012; Henry et al., 2019). This process yields a sparse, person-specific network for each participant that includes only connections that are meaningful to model fit, with some shared across the full sample, some only within a subgroup (EF-CBT, RMT, L/NA), and some unique to an individual. CS-GIMME does not average connectivity estimates across participants.

Because the resulting networks vary in the connections estimated across individuals, inference was conducted using network-level summary metrics rather than edgewise comparisons, following others (Becker et al., 2024; Beltz & Gates, 2017; Hardi et al., 2024). We focused on three density metrics: within-CEN, CEN-SN, and CEN-DMN. For each participant and timepoint, network density was defined as the proportion of contemporaneous and lagged connections within or between the relevant networks, compared to the total number of estimated connections between all ROIs in the sparse network.

### Statistical Analyses

<u>Clinical</u>: Clinical analyses tested changes in PARS scores from pre- to post-treatment using linear mixed-effects (LME) models, with comparisons conducted within and between treatment groups. CGI-I scores were also compared between treatment groups using an independent *t*-test.

<u>Connectivity</u>: First, linear regression models compared baseline differences in network density between patients and L/NA participants. Next, treatment-related change was assessed with LME models, with the interaction of treatment condition (EF-CBT, RMT) and time (baseline, follow-up) nested within participant, on each of the three connectivity metrics. These models included age and in-scanner motion (FD) as covariates, and results were corrected for multiple comparisons using the false discovery rate (FDR; Benjamini & Hochberg, 1995). To examine change from baseline to follow-up, significant time-by-treatment interactions were probed for each treatment group with paired *t*-tests without covariates on complete case data. Changes in these connectivity metrics were tested for associations with change in symptoms, within treatment group. Additionally, post hoc LME models evaluated changes in network density from baseline to follow-up in L/NA participants, controlling for age and FD. Finally, post hoc analyses examined the association between change in network density and symptom change in patients. This analysis tested the interacting effect of density change and treatment group on post-treatment PARS in the complete case sample, controlling for baseline PARS, age, and average FD.

## Results

### Demographic and clinical characteristics

One-hundred and four anxious youth and 37 L/NA youth had usable resting-state scans at baseline. Of the 104 patients, 70 received EF-CBT and 34 received RMT. At the follow-up scan approximately 16 weeks later, 113 participants (including 34 L/NA youth) had usable resting-state data. A total of 97 participants (48 EF-CBT, 21 RMT, 28 L/NA) had complete resting-state data at both timepoints (i.e., “complete cases”). Baseline demographic and clinical characteristics by treatment and diagnostic condition are reported in Table 1. See Figure S1 for a CONSORT diagram.

**Table 1.** Baseline Demographic and Clinical Characteristics.

|  | **ANX** | **L/NA** | **EF-CBT** | **RMT** |
| --- | --- | --- | --- | --- |
|  | *N* = 104 | *N* = 37 | *N* = 70 | *N* = 34 |
|  | *M* (*SD*) | *M* (*SD*) | *M* (*SD*) | *M* (*SD*) |
| **Demographic** |  |  |  |  |
| Age (years) | 12.22 (2.98) | 13.03 (3.20) | 11.90 (3.09) | 12.88 (2.66) |
| Sex |  |  |  |  |
| Female | 83 | 20 | 56 | 27 |
| Male | 21 | 17 | 14 | 7 |
| Race |  |  |  |  |
| Asian | 4 | 3 | 3 | 1 |
| Black/African American | 4 | 2 | 0 | 4 |
| Multiracial | 13 | 6 | 9 | 4 |
| Other Pacific Islander | 1 | 0 | 0 | 1 |
| White | 81 | 25 | 58 | 23 |
| Unreported/unknown | 1 | 1 | 0 | 1 |
| Ethnicity |  |  |  |  |
| Non-Hispanic/Latinx | 101 | 35 | 68 | 33 |
| Hispanic/Latinx | 3 | 2 | 2 | 1 |
| **Baseline clinical** |  |  |  |  |
| PARS | 19.60 (3.29) | 6.05 (3.69) | 19.43 (3.10) | 19.66 (3.66) |
| MASC | 60.00 (10.49) | 40.43 (10.09) | 59.37 (10.18) | 61.18 (11.10) |
| CDI | 12.75 (7.83) | 2.59 (2.73) | 12.58 (7.98) | 13.13 (7.60) |

*Note.* M = Mean; SD = Standard deviation; EF-CBT = Exposure-focused cognitive behavioral therapy; RMT = relaxation mentorship training; ANX = group of youth with diagnosed anxiety; L/NA = low/no anxiety group; PARS = Pediatric Anxiety Rating Scale; MASC = Multidimensional Anxiety Scale for Children; CDI = Children's Depression Inventory.

Results of the full clinical trial, which included participants with missing or unusable resting-state data, have been previously reported and found greater anxiety symptom reduction in patients randomized to EF-CBT compared to RMT on both the PARS and CGI-I (Bilek et al., 2021, 2025). In the present imaging subsample, significant pre-to-post-treatment reductions in PARS scores were observed following both EF-CBT (*b* = −5.56, *t*(58.1) = −9.79, *p* < .001) and RMT (*b* = −4.07, *t*(23.7) = −5.07, *p* < .001), but symptom change did not significantly differ by treatment group (*b* = −1.47, *t*(82.5) = −1.46, *p* = .15). However, improvement on the CGI-I was greater for patients who received EF-CBT (M_CGI-I_ = 2.35) than RMT (M_CGI-I_ = 3.30), *t*(94) = 5.18, *p* < .001.

### Characteristics of CS-GIMME models

All CS-GIMME-derived models met individual-level stopping rules requiring ≥ 2 of 4 stringent fit thresholds for all individual-level networks. At baseline (N = 141), there were a total of 21 full-sample connections, as well as subgroup-specific connections (EF-CBT: 3; RMT: 9; L/NA: 12). At follow-up (N = 113), there were 23 full-sample connections, as well as subgroup-specific connections (EF-CBT: 2; RMT: 5; L/NA: 6). Beyond these shared structures, participants also showed person-specific connections, consistent with CS-GIMME’s design to capture both common and individual features of network organization (Figure S2). Additional model characteristics are reported in the Supporting Information.

### Baseline differences in network density

At baseline, anxious youth had weaker CEN-SN density (*b* = −.037, *t*(137) = −6.37, *p* < .001), CEN-DMN density (*b* = −.062, *t*(137) = −11.37, *p* < .001) and within-CEN density (*b =* −.048, *t*(137) = −10.06, *p* < .001) than L/NA youth (*Figure 1*).

**Figure 1.**
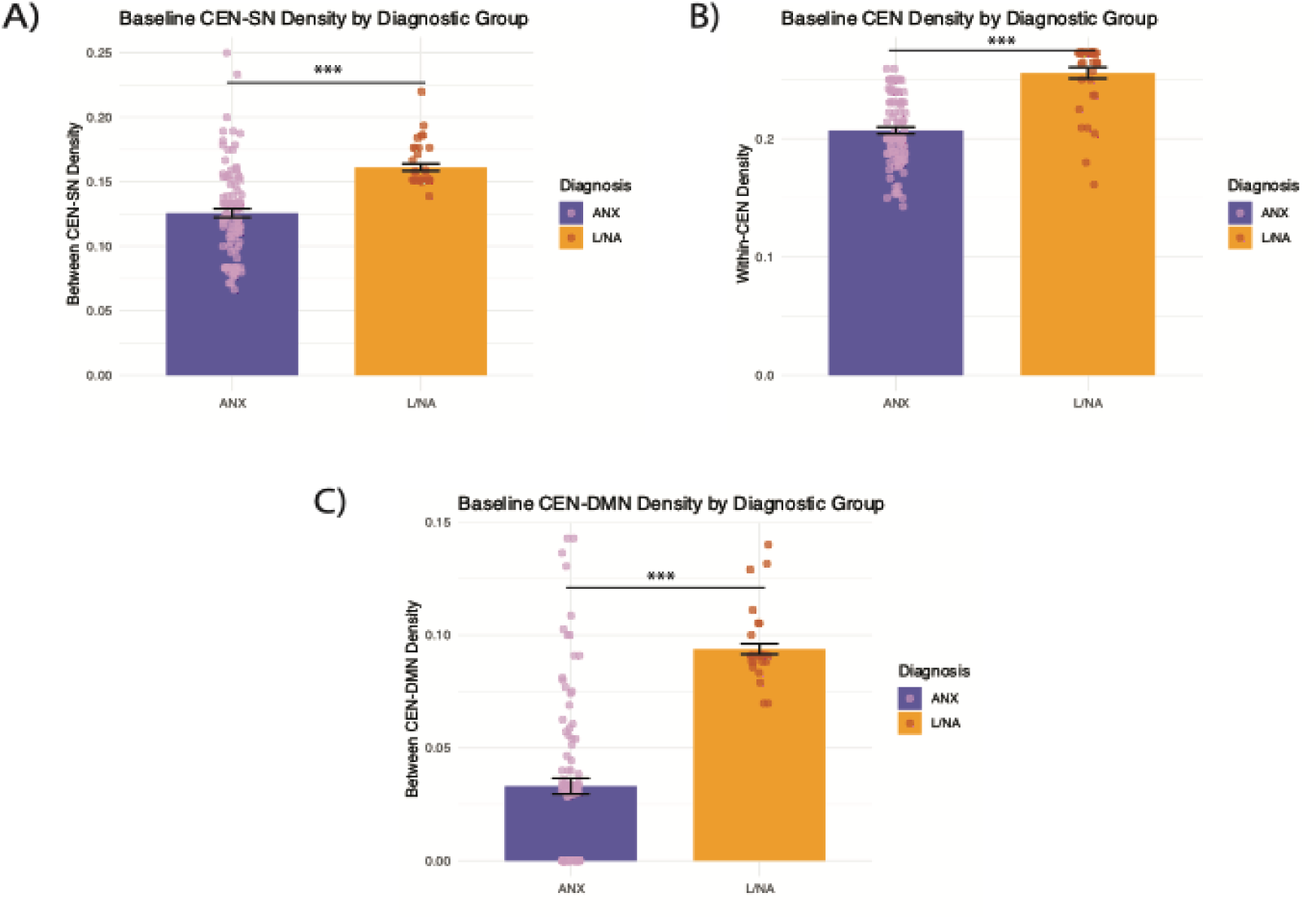
**Baseline network density by diagnostic group.** *Note.* Baseline differences in A) CEN-SN density, B) Within-CEN density, and C) CEN-DMN density between patients with anxiety disorders (ANX) and low/no anxiety youth (L/NA). Bars reflect mean density and error bars indicate standard error. The dots on the bars reflect density values for each participant. CEN = Central Executive Network; SN = Salience Network; DMN = Default Mode Network. \*\*\**p* < .001.

### Treatment-related changes in network density

In anxious youth, time and treatment condition interacted to predict CEN-SN density (*b* = 0.08, *t*(101.25) = 7.41, *p*FDR < .001, 95% CI[0.06, 0.10]) and within-CEN density (*b* = −.05, *t*(103.86) = −5.63, *p*FDR < .001, 95% CI[-0.06,- 0.03]) (*Figures 2 and 3; Table 2*). EF-CBT led to a significant increase in CEN-SN density (*t*(47) = −4.89, *p* < .001), whereas density decreased after RMT (*t*(20) = 5.04, *p* < .001). By contrast, within-CEN density increased from pre- to post-RMT (*t*(20) = −7.30, *p* < .001), but did not significantly change following EF-CBT (*t*(47) = 1.05, *p* = 0.30). No time-by-treatment interaction emerged for CEN-DMN density (*b* = −0.0002, *t*(93.11) = −.019, *p*FDR = .99, 95% CI[-0.02, 0.02]). No significant associations were observed between change in anxiety symptoms and change in CEN-SN density or within-CEN density (see Supporting Information). In L/NA participants, there was a main effect of time for CEN-SN density (*b* = −.071*, t*(38.68) = −13.94*, p <* .001), such that CEN-SN density significantly decreased from baseline to follow-up. L/NA youth did not experience changes in within-CEN density (*b* = −.002, *t*(36.21) = −.62, *p* = .54). Analyses comparing change in the L/NA group to change in the treatment groups are reported in the Supporting Information (*Table S2*; *Figure S3*).

**Figure 2.**
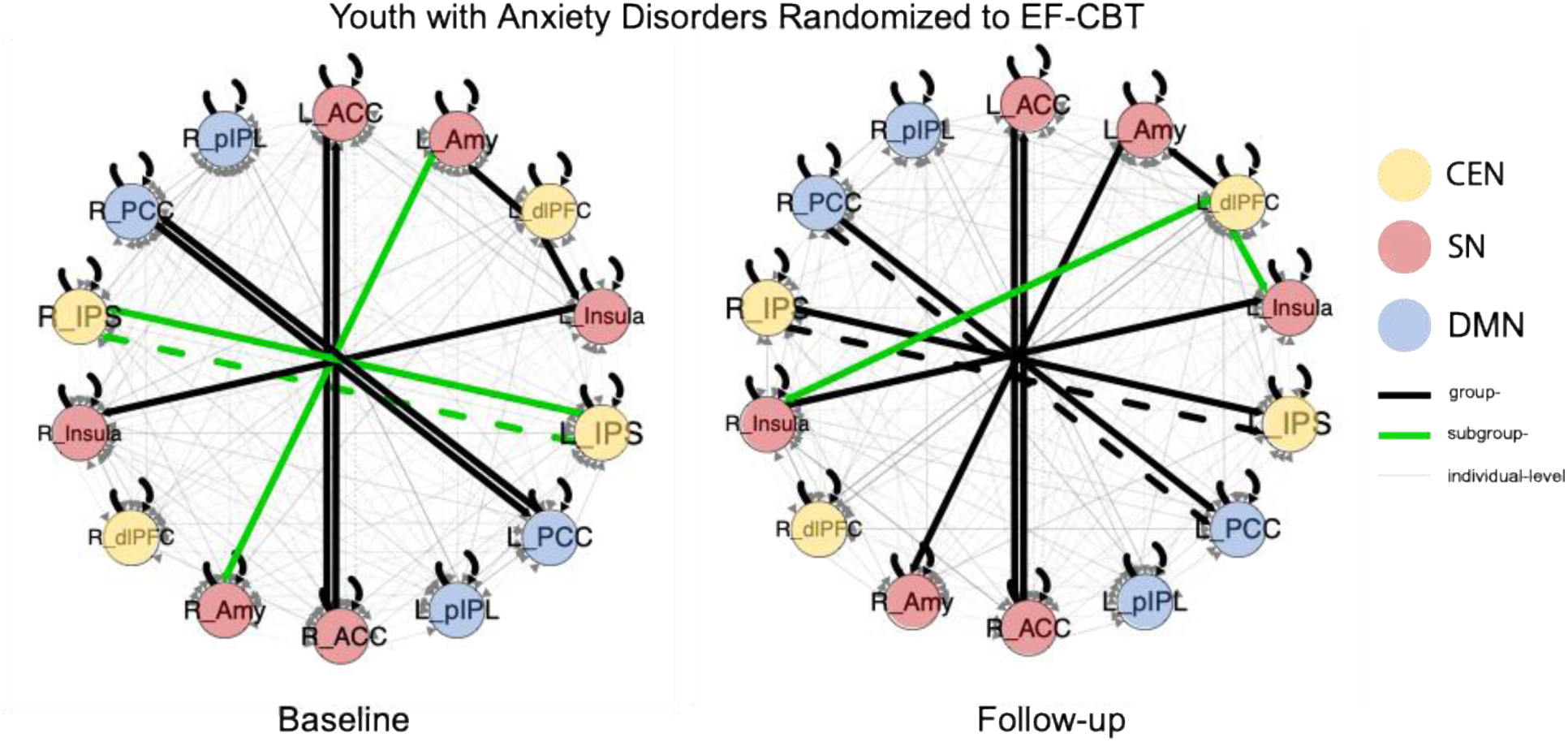
**Summary of person-specific connectivity maps before and after EF-CBT.** *Note.* Changes in network density in anxious youth following exposure-focused CBT (EF-CBT). Black lines indicate group-level connections, green lines indicate connections specific to EF-CBT, and gray lines indicate individual connections. Line thickness reflects the proportion of participants for whom the connection was estimated. Solid lines indicate contemporaneous connections and dashed lines indicate lagged connections. CEN = central executive network; SN = salience network; DMN = default mode network. R/L = right/left; ACC = anterior cingulate cortex; Amyg = amygdala; dlPFC = dorsolateral prefrontal cortex; Ins = insula; IPS = intraparietal sulcus; PCC = posterior cingulate cortex; pIPL = posterior inferior parietal lobule.

**Figure 3.**
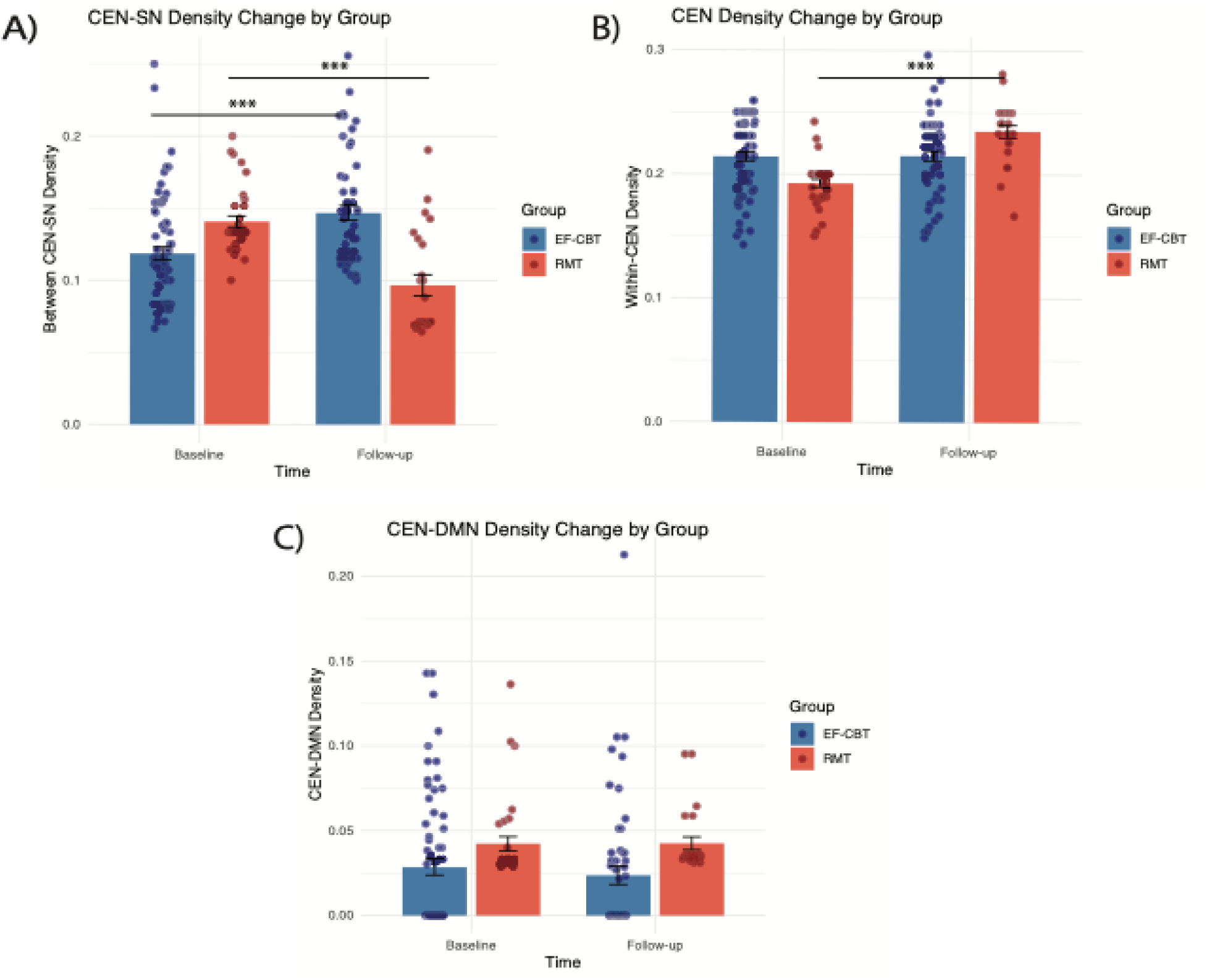
**Change in network density over time by treatment condition.** *Note.* Pre-to-post treatment change in density A) between CEN-SN, B) within-CEN, and C) between CEN-DMN by treatment condition. Bars represent means with standard error, and dots represent individual participants. CEN = central executive network; SN = salience network; DMN = default mode network; EF-CBT = exposure-focused cognitive behavioral therapy; RMT = relaxation mentorship training. *** *p* < .001.

**Table 2.** LME model results for pre-to-post-treatment changes in network density.

| **CEN-SN Density** |  |  |  |  |  |
| --- | --- | --- | --- | --- | --- |
|  | *Estimate* | *SE* | *df* | *t* | *p* |
| Intercept | 0.16 | 0.01 | 147.65 | 10.98 | <.001 |
| Treatment | -0.03 | 0.01 | 177.00 | -4.00 | <.001 |
| Time | -0.04 | 0.01 | 102.30 | -5.14 | <.001 |
| Treatment x Time | 0.08 | 0.01 | 101.25 | 7.41 | <.001 |
| Age | 0.00 | 0.00 | 122.45 | -1.96 | 0.05 |
| Framewise Displacement | 0.05 | 0.01 | 177.00 | 3.19 | 0.00 |

| **Within-CEN Density** |  |  |  |  |  |
| --- | --- | --- | --- | --- | --- |
|  | *Estimate* | *SE* | *df* | *t* | *p* |
| Intercept | 0.20 | 0.01 | 147.42 | 18.33 | <.001 |
| Treatment | 0.03 | 0.01 | 177.00 | 5.05 | <.001 |
| Time | 0.04 | 0.01 | 104.95 | 6.29 | <.001 |
| Treatment x Time | -0.04 | 0.01 | 103.86 | -5.63 | <.001 |
| Age | 0.00 | 0.00 | 121.84 | 0.69 | 0.49 |
| Framewise Displacement | -0.06 | 0.01 | 177.00 | -5.80 | <.001 |

| **CEN-DMN Density** |  |  |  |  |  |
| --- | --- | --- | --- | --- | --- |
|  | *Estimate* | *SE* | *df* | *t* | *p* |
| Intercept | 0.04 | 0.01 | 146.14 | 3.31 | 0.001 |
| Treatment | -0.02 | 0.01 | 177.00 | -3.38 | <.001 |
| Time | 0.00 | 0.01 | 94.07 | 0.18 | 0.86 |
| Treatment x Time | -0.00 | 0.01 | 93.11 | -0.02 | 0.99 |
| Age | -0.00 | 0.00 | 121.02 | -1.43 | 0.15 |
| Framewise Displacement | 0.08 | 0.01 | 177.00 | 6.34 | <.001 |

*Note.* EF-CBT = exposure-focused CBT; RMT = relaxation mentorship training; CEN = central executive network; SN = salience network; DMN = default mode network; LME = linear mixed effects.

### Connectivity change association with symptom change

There were no significant associations between symptom change and change in CEN-SN or within-CEN density in the complete case sample of anxious youth (CEN-SN: *p* = .17; within-CEN: *p* = .45), nor did network density interact with treatment group to predict symptoms (CEN-SN: *p* = .30; within-CEN: *p* = .52).

## Discussion

This RCT of pediatric anxiety tested within and between network density of the CEN with the SN and DMN relative to youth without clinical anxiety, and examined treatment-related change with EF-CBT versus RMT. As hypothesized, anxious youth displayed reduced within-CEN and between CEN-SN density at baseline, suggesting attenuated coordination of core cognitive control and salience-control systems relative to L/NA youth. Furthermore, EF-CBT selectively increased CEN–SN density, whereas RMT did not, consistent with our hypothesis that exposure may increase neural capacity for cognitive control (Fitzgerald et al., 2021). Contrary to our hypotheses, anxious youth showed reduced rather than greater CEN-DMN connectivity than L/NA youth, and EF-CBT did not lead to significant changes in within-CEN or CEN-DMN density. Given the heterogeneity of pediatric anxiety across development, employing GIMME for person-specific estimation offered a distinct advantage over group-averaging, enhancing our capacity to detect treatment-related changes in network organization. Together, these findings suggest that disrupted cognitive control network (CEN) organization and weakened control-salience integration may underlie pediatric anxiety, and that exposure therapy strengthens coordination between these systems.

### Lower within-CEN and CEN-SN density in anxious youth

In line with our hypotheses, anxious youth displayed lower person-specific network density within-CEN and between CEN-SN at baseline than L/NA youth. This is consistent with tripartite network models of anxiety, in which weakened intrinsic coordination of cognitive control circuitry and reduced coupling between control and salience systems compromise goal-directed regulation of worry-driven attention and avoidance behavior (Fitzgerald et al., 2021; Menon & D’Esposito, 2022; Sylvester et al., 2012). Weaker within-CEN and CEN-SN connectivity has been associated with cognitive control deficits (Cai et al., 2021; Cole & Schneider, 2007; Dosenbach et al., 2006; Menon & D’Esposito, 2022), supporting the hypothesis of reduced neural capacity for cognitive control in patients with anxiety disorders (Sylvester et al., 2012). Furthermore, resting-state studies have linked elevated anxiety to reduced executive–salience coupling, including in trait-anxious adolescents and in adult anxiety samples (Geng et al., 2016, Xu et al., 2019). Extending this work, the present findings of lower person-specific network density within-CEN and between CEN-SN across a broad age range suggest that pediatric anxiety is characterized by weakened intrinsic control architecture and diminished integration of cognitive control with salience detection, potentially limiting the flexible recruitment of control processes in response to salient or threatening stimuli.

### Normalization of CEN-SN density following EF-CBT

Between CEN-SN density selectively increased (i.e., normalized) in patients who received EF-CBT, consistent with our hypothesis that exposure engages neural interactions supporting cognitive control. In the present analyses, the amygdala was included as an SN node given its central role in detecting motivationally salient and threat-related information and in signaling the need for control (Swartz & Monk, 2013), and its well-established hyperactivity in youth anxiety (e.g., Ashworth et al., 2021; Díaz et al., 2024; Monk et al., 2008). Thus, exposure may enhance top-down modulation of salience signals from the amygdala, and the SN more broadly (Hauner et al., 2012). Consistent with this interpretation, prior work shows that pretreatment activation in key CEN and SN regions during emotionally salient tasks predicts CBT response in anxious youth (Burkhouse et al., 2017; Díaz et al., 2024; Diaz et al., 2025; Kujawa et al., 2016), and CEN activity normalizes following CBT (Haller et al., 2024). Interestingly, both L/NA youth and anxious youth who underwent RMT exhibited decreases in CEN–SN density. This may reflect normative habituation and reduced salience across repeated scans, or other nonspecific effects of time, which may occur when CEN-SN coupling is not practiced through repeated exposure. Together, these findings suggest that exposure-focused CBT uniquely strengthens control–salience network integration, identifying CEN–SN density as a modifiable neural mechanism of treatment response and a potential target for pre-treatment interventions to optimize exposure efficacy by enhancing cognitive control.

### Within-CEN density normalizes with RMT

Contrary to our hypothesis, person-specific network density within the CEN did not increase following EF-CBT, and instead increased with RMT. This pattern suggests that EF-CBT may act more selectively on CEN control over salience processing rather than intrinsic CEN integration, consistent with exposure’s emphasis on engaging cognitive control to tolerate feared stimuli without avoidance. Notably, GIMME estimates sparse networks by prioritizing connections that best explain system dynamics. Thus, the absence of significant within-CEN change after EF-CBT does not necessarily signal that the network was unaffected; it may instead indicate that other pathways (e.g., CEN–SN coupling) better captured treatment-related reorganization.

By contrast, within-CEN density increased following RMT. Although unanticipated, this finding aligns with prior work on mindfulness-based interventions, which similarly emphasize sustained, internally focused attention. Mindfulness training has been associated with increased CEN connectivity in children with anxiety-related attention impairments (Kennedy et al., 2022), and adults with elevated psychological distress (Taren et al., 2017). Although distinct interventions, both mindfulness training and RMT engage tonic control processes (e.g., maintaining attention to breath or bodily sensations and re-engaging attention after distraction), which may strengthen CEN integration. Together, these findings indicate that exposure and relaxation exert dissociable effects on cognitive control network organization, reflecting distinct forms of cognitive control plasticity and underscoring the value of incorporating both components within CBT. Future work should examine whether CBT can be optimized by tailoring subcomponents to individual network function.

### Lower CEN-DMN density in anxiety

Network models of anxiety propose poor segregation and inefficient switching between the DMN and task-control networks, largely based on evidence of task-evoked DMN hyperactivation and impaired suppression of perseverative internal thought (Fitzgerald et al., 2021; Menon & D’Esposito, 2022). On that basis, we predicted CEN–DMN density would be higher for anxious than L/NA youth, reflecting difficulty controlling repetitive negative worry, and that it would normalize with exposure practice. Instead, anxious youth showed lower CEN–DMN density than L/NA youth, with no change following EF-CBT or RMT. Although DMN alterations are well documented in anxiety, the direction of intrinsic connectivity effects is inconsistent, both within the DMN (Kim & Yoon, 2018; Li et al., 2023) and between the DMN and task-positive networks (Luo et al., 2025; Xu et al., 2019). One of the few reports implicating CEN–DMN alterations directly is a meta-analysis across clinical and trait anxiety showing *reduced* connectivity between dorsolateral prefrontal cortex and DMN regions (Xu et al., 2019), in line with our findings. Thus, anxiety may reflect under-engagement of CEN–DMN coupling, rather than excessive or maladaptive integration. That is, failed suppression of perseverative thought may stem not from intrusive control of the DMN, but from insufficient recruitment of the CEN to modulate DMN activity, potentially indicating a failure to initiate control rather than failure to implement it. However, CEN-DMN did not change with treatment, suggesting this network connection is less relevant to exposure efficacy than other CEN density metrics.

### Limitations

Several limitations should be noted. First, although large for an fMRI RCT of treatment mechanisms, the study sample was smaller than CBT efficacy trials powered to detect anxiety symptom change on the PARS (e.g., CAMS; Walkup et al., 2008), due to unequal treatment group sizes and conservative motion thresholds. Nonetheless, we observed better symptom improvement on the CGI-I following EF-CBT than RMT, and analyses of the full clinical trial sample from this study demonstrated significant treatment effects on PARS scores (Bilek et al., 2021, 2025). Second, although changes in intrinsic network connectivity differed by treatment type, they were not associated with symptom improvement. Changes in intrinsic connectivity may reflect engagement of control processes that support learning during treatment, whereas symptom reduction is a downstream and heterogeneous outcome that may not map linearly onto pre–post neural change. This study was powered to detect treatment effects on neural connectivity using a within-subject fMRI design (Lee et al., 2024), but future studies with larger sample sizes may be better positioned to detect associations between neural change and symptom improvement. Third, the study sample was predominantly female, which may limit the generalizability of these findings to males, but is consistent with work showing anxiety disorders are at least twice as common in females (Lewinsohn et al., 1998). Finally, GIMME relies on *a priori* ROI selection. This aligned with our focus on CEN-related connectivity, but limited inferences about connectivity across other brain regions, as different networks or ROIs selections may yield distinct findings (Henry et al., 2019). At the same time, GIMME’s ability to estimate group-, subgroup-, and person-specific network structure increases robustness to heterogeneity, which is particularly important in pediatric clinical samples.

## Conclusion

In this randomized clinical trial, pediatric anxiety was characterized by reduced intrinsic connectivity of tripartite networks and weaker coordination between control (CEN) and salience (SN) systems at rest. Exposure-focused CBT selectively increased CEN–SN density, whereas relaxation training increased within-CEN density, indicating dissociable neural mechanisms of treatment effect. These findings suggest that exposure uniquely strengthens control–salience integration, identifying a network-level mechanism that may be leveraged to optimize or personalize intervention strategies in pediatric anxiety.

## Data Availability

The data that support the findings of this study are available from the corresponding author upon reasonable request.

## Acknowledgements

This project was supported by the National Institutes of Health (R01 MH107419), National Institute of Child Health and Human Development (T32 HD007109). The authors would also like to thank the children and families who participated in the study.

## Ethics Statement

Participants and guardians provided informed assent/written consent to participate, in compliance with the Michigan Medicine Institutional Review Board (IRB; HUM00118950). The original date of IRB Approval was 7/29/2016.

## Declaration of interests

Authors have no disclosures.

## CRediT statement

Hannah Becker: conceptualization, formal analysis, writing - original draft; Adriene Beltz: methodology, writing - review; Emily Bilek: writing - review; Dana Diaz: writing - drafting and review; Kate Fitzgerald: conceptualization, funding acquisition, supervision, writing - review; Felicia Hardi: conceptualization, formal analysis, writing - review; Chris Monk: conceptualization, funding acquisition, supervision, writing - review; Luan Phan: funding acquisition, writing - review; Stefanie Russman Block: writing - review.

## Notes

### Competing Interest Statement

The authors have declared no competing interest.

### Clinical Trial

Clinical Trial Number: NCT02810171.

### Clinical Protocols

https://clinicaltrials.gov/study/NCT02810171

