## Supplementary Material for "Connectivity between the central executive and salience networks normalizes with exposure-focused CBT in pediatric anxiety"

**Supporting Information**

**Supporting Methods**

**Demographic and clinical assessment**

***Demographic questionnaire***

Demographic information, including biological sex, race, and ethnicity, was collected using a demographic questionnaire that was developed for the purposes of this study. Each participant’s caregiver completed this form on behalf of each youth participant.

***Pediatric Anxiety Rating Scale***

The Pediatric Anxiety Rating Scale (PARS; Group, 2002) is a semi-structured clinical interview that was used to assess anxiety severity dimensionally in all groups. Overall PARS severity scores are derived from impairment scores associated with a list of anxiety symptoms spanning different domains of anxiety (e.g., separation anxieties, social anxieties, physical symptoms of anxiety). Higher PARS scores indicate greater anxiety severity (score range: 0-30). Youth and their guardian(s) were interviewed together or separately for the PARS assessment, though almost all families chose to be interviewed with child and guardian together. The clinical rater was an independent evaluator (i.e., masked to treatment condition). The PARS is a common outcome measure in treatment studies for anxiety (Walkup et al., 2008). Anxiety symptoms were assessed with the PARS at the first, sixth, ninth, and final treatment sessions. Additionally, depression symptoms were assessed with the Children’s Depression Inventory (CDI) and overall symptom improvement was assessed with the Clinical Global Impression-Improvement (CGI-I) scale.

***Children’s Depression Inventory (CDI)***

The CDI (Kovacs, 1978) is a self-report measure of depressive symptoms. All participants completed this questionnaire at baseline and follow-up. Higher scores indicate greater levels of depressive symptoms (score range: 0-54). The CDI was included in the current study to understand the potential effects of depressive symptom comorbidity on anxiety disorders in youth and change with treatment.

***Clinical Global Impression–Global Improvement (CGI-I)***

The CGI-I (Guy, 1976) is a clinician-rated measure of patient improvement since the baseline assessment. Scores from the CGI-I assessment at follow-up (~week 12), conducted by an independent evaluator, were used in the current analyses. Scores on the CGI-I range from 1-7, with lower scores reflecting greater improvement since the start of treatment. A score of 1 indicates that the patient’s symptoms were “very much improved.”

**Resting-state fMRI acquisition and preprocessing**

Whole-brain blood-oxygen-level dependent (BOLD) fMRI data were collected using multiband imaging. The resting-state functional images were acquired on a 3.0 Tesla GE Discovery MR750 scanners using multiband gradient-echo EPI (TR=800 ms, TE=30 ms, flip angle (FA)=52°, multiband acceleration factor = 6, 2.4-mm isotropic voxels, 60 slices, FOV=216 mm). In order to facilitate co-registration with the functional images, a low-resolution axial T1 was collected. Additionally, high-resolution T1 weighted scans were acquired with 3D MPRAGE sequence (TR=2500 ms, TE=1.9 ms, TI=1060 ms, FA=8°,1.0-mm isotropic voxels, 176 slices, FOV=256 mm) for the normalization of the functional images. Participants were instructed to remain awake and still for the duration of the scan and focus on the fixation cross in the center of the screen. Head movement was further minimized through packing with foam padding.

Anatomical images were skull-stripped (f=.25) using the Brain Extraction Tool (BET) in FSL version 6.0 (Jenkinson et al., 2012) and segmented into gray matter, white matter, and cerebrospinal fluid using FSL FAST. The first ten volumes of functional data were removed to ensure the stability of signal intensity. Then, functional images were skull-stripped and spatially smoothed using FSL and registered to subject-specific previously skull-stripped and segmented anatomical images. A temporal derivative was applied, as well as motion correction (using MCFLIRT) and spatial smoothing, using a Gaussian kernel of FWHM 6.0mm. ICA-AROMA (Pruim et al., 2015) was used to remove motion-related artifacts in the data, nuisance signal derived from white matter and cerebrospinal fluid were regressed out, and data with signal below 0.01Hz were then high-pass filtered. These preprocessing steps were implemented using scripts (Beltz et al., 2019) that were also utilized in previous investigations (Goetschius et al., 2020; Hardi et al., 2023). Mean framewise displacement for each subject, at each timepoint, was calculated by averaging the mean framewise displacement (FD; Power et al., 2012) from both resting-state runs at each timepoint. The NeuroSynth coordinates selected for the ROIs were also cross-referenced qualitatively with coordinates from the Seitzman atlas (Seitzman et al., 2020).

Anxiety symptoms (as measured by the PARS), framewise displacement, and treatment group membership were summarized in the sample of excluded participants who had baseline or follow-up resting-state fMRI data, separately. These characteristics were statistically compared to those of the sample of included participants using independent *t*-tests. Excluded participants who had resting-state fMRI data were also separated into groups based on exclusion reason (high FD or failed preprocessing, etc.) and compared to the included group. These results are reported in the Supplemental Results. Of note, follow-up fMRI data from one participant (assigned to exposure-focused cognitive behavioral therapy; EF-CBT) with only one run of follow-up data was accidentally included in the follow-up GIMME model, though this participant converged normally in the model and did not impact the results in any observable way.

**GIMME process and rationale: additional detail**

Connections can be in multiple subgroups (i.e., not limited to a specific subgroup) and connections can belong to both a subgroup and an individual, depending on the individual. An additional strength of GIMME is that it estimates time-locked (contemporaneous) and lagged positive and negative directed connections *between* regions. Both positive and negative connections are included in the path estimation and subsequent density calculations, though the positive or negative valence of the density was not a focus of the current analyses. Still, sensitivity analyses were conducted to better understand the proportion of positive and negative connections in each density metric. The number of positive connections were summed for each subject’s specific density metric and divided by the total number of connections (positive and negative) for that subject’s density metric (i.e., for each subject: N of positive within-CEN connections / total N of within-CEN connections).

**Statistical analyses**

***Weeks of treatment and number of treatment sessions by treatment group***

Descriptive summary statistics (means and standard deviations) were calculated for treatment duration (number of weeks) and number of treatment sessions for EF-CBT and RMT groups. Independent t-tests tested for differences on these measures between EF-CBT and RMT groups.

***Differences in connectivity change between EF-CBT, RMT, and low/no anxiety groups over time***

Post-hoc LME models were run to directly compare the change in the L/NA group to the other two groups (EF-CBT and RMT). These models assessed the interacting effect of time and group on density, with “group” treated as a 3-level categorical variable, and the L/NA group as the reference group. As with the LME models assessing change in density over time in patients, in-scanner motion and age were included as covariates.

***Association between baseline density and treatment response***

Post-hoc analyses were conducted to better understand how baseline network density related to eventual treatment outcomes (i.e., anxiety symptom reduction). These analyses used linear mixed effects models to examine the effect of baseline density metrics (CEN, CEN-SN, and CEN-DMN density) on follow-up PARS scores, controlling for baseline PARS scores. These analyses were conducted with and without an interaction of treatment (EF-CBT or RMT), separately within each treatment group, and were conducted in both the full usable baseline sample as well as the complete case sample. Complete case analyses were favored for this analysis because each participant with a baseline density metric also had a follow-up PARS score, though, results from both samples are reported. Age and mean FD were included as covariates each of these models. Given, the post hoc nature of these analyses, results were examined without correction for multiple comparisons.

**Supporting Results**

**Weeks of treatment and number of treatment sessions by treatment group**

In the full sample of patients with usable pre-treatment fMRI data (N = 104), participants were engaged for an average of 13.12 (SD = 2.06) weeks of treatment. The number weeks varied by treatment received, such that participants randomized to EF-CBT received treatment for an average of 13.55 (SD = 1.76) weeks and those in RMT received an average of 12.18 weeks (SD = 2.37) of treatment, *t*(44.25) = -2.85, *p* = .007. The average number of sessions completed across groups was 10.87 (SD = 2.52) sessions. Again, this varied by treatment group such that participants randomized to EF-CBT completed an average of 11.29 (SD = 2.00) sessions, and those randomized to RMT completed an average of 10.00 (SD = 3.22) sessions, *t*(45.78) = -2.14, *p* = .038. This information is also reported in a previous clinical investigation of this clinical trial (Bilek et al., 2021).

**Characteristics of fMRI sample excluded for motion or preprocessing failure**

There was no significant difference in PARS scores (anxiety symptoms) between those excluded and those included at baseline, nor at follow-up. At baseline, there was a significant difference between those excluded and included in terms of age, such that those who were excluded were significantly younger than those included, *p* = .014. This was driven by participants excluded for high FD (*p* = .002), and not those excluded for other reasons such as failing preprocessing, post-preprocessing QC, or only having one run (*p* = .73). At follow-up, there was again a significant difference in age between those excluded than those included, (*p* = .019), which was again driven by those excluded for high FD (as compared to those included at follow-up: *p* < .001), rather than other reasons for exclusion (*p* = .45). Finally, the proportion of participant scans excluded at baseline and follow-up in terms of treatment assignment to EF-CBT was 68.5% and 70.3%, respectively, which is in alignment with the 2:1 randomization ratio. In other words, a proportionally comparable amount of participants were excluded from each treatment group, at each timepoint.

**Additional CS-GIMME Model Characteristics**

Across participants, there were an average of 4.84 (SD = 6.28) individual-specific connections in the baseline model. In the follow-up model, there were an average of 4.63 (SD = 5.59) individual-specific connections. Follow-up analyses were conducted to better characterize the proportion of positive and negative connections in each subject’s density metrics of within-CEN, CEN-SN, and CEN-DMN density. At baseline, the average (across subjects) proportion of positive connections was 79% for within-CEN density, 58% CEN-SN density, and 45% CEN-DMN density. Therefore, the relative majority of connections were positive for baseline within-CEN and CEN-SN density, and the relative majority were negative for baseline CEN-DMN density. The proportion of positive connections remained consistent at follow-up: 81% for within-CEN density (more positive), 58% for CEN-SN density (more positive), and 45% for CEN-DMN density (more negative connections).

**Differences in connectivity change between EF-CBT, RMT and low/no anxiety groups over time**

Results for this analysis are presented in Supporting Table 1. In summary, for CEN-SN density, there were significant time by treatment interactions for EF-CBT compared to L/NA, indicating an increase in density over time for EF-CBT. A time by treatment interaction was also significant for RMT compared to L/NA; both groups exhibited the same pattern of decreasing CEN-SN density, but density was lower at baseline in the RMT group as compared to the L/NA group. For within-CEN density, there was a significant time by treatment interaction only for RMT compared to L/NA, indicating an increase in density for RMT and no change for the L/NA group, whereas there was no difference between EF-CBT and L/NA groups in within-CEN density change over time.

**Baseline connectivity association with treatment response**

In the full sample with usable baseline data, there were no significant interactions between baseline density and treatment on follow-up PARS score, for any of the three density metrics, all *p* > .492. Additionally, baseline density also did not significantly associate with symptoms at follow-up in models that examined change across both treatments (i.e., independent of treatment received), all *p >* .090. However, within- treatment group analyses revealed a significant effect: patients randomized to EF-CBT who had higher CEN-SN density at baseline had lower symptoms at follow-up, *b*=-26.20, *t*(61)=-2.15, *p*=.036.

**Sensitivity analyses in complete case dataset**

*The complete case dataset includes participants who had usable fMRI data at both baseline and follow-up (N_Total_ = 97; N_CBT_ = 48 ; N_RMT_ = 21; N_L/NA_ = 28). The full sample included participants with usable fMRI data at baseline or follow-up, resulting in a larger sample, but included some participants that only had data from one timepoint (either baseline or follow-up). Given that the nature of this study was to evaluate baseline to follow-up changes that occurred with treatment, sensitivity analyses in smaller, complete case dataset are included here that evaluate change over time in participants who had completed both timepoints (i.e., before and after treatment).*

***Clinical data*** ***(complete case)***

Significant reduction in PARS scores were observed in both EF-CBT (*t*(47)=9.68, *p*<.001, d=1.59) and RMT groups (*t*(20)=4.70, *p*<.001, d=0.81) from baseline to follow-up, but reductions did not differ by treatment group (*b*=-1.86, *t*(67)=-1.75, *p*=.085). Clinical Global Impressions-Improvement (CGI-I) scores indicated significantly greater improvement in patients who received EF-CBT (M_CGI-I_ :2.35) than RMT (M_CGI-I_: 3.19), *t*(67)=3.69, *p*<.001. These results were consistent with changes observed in the full usable sample (reported in manuscript text).

***Baseline differences in patients compared to low/no anxious youth (complete case)***

At baseline, patients with anxiety had weaker CEN-SN density (*b*=-.037, *t*(93)=-5.86, *p*<.001), CEN-DMN density (*b*=-.064, *t*(93)=-11.96, *p*<.001) and within-CEN density (*b*=-.045, *t*(93)=-8.33, *p*<.001) than L/NA youth, reflecting different starting points in neural substrate for cognitive control. These results are consistent with those in the full usable sample.

***Baseline to follow-up brain change (complete case)***

In patients, significant interactions between time and treatment condition were observed for CEN-SN density (*b*=0.077, *t*(69.2)=6.71, *p*<.001, *p*FDR<.001, 95% CI[0.051,0.10]) and within-CEN density (*b*=-.052, *t*(69.2)=-5.76, *p*<.001, *p*FDR<.001, 95% CI[-0.070,-.034]. There was no significant interaction between time and treatment condition for CEN-DMN density (*b*=-2_x_10^-5^, *t*(69.2)=.002, *p*=.998, *p*FDR=.998, 95% CI[-0.019,0.019]). In the L/NA participants, there was a significant main effect of time for CEN-SN density (*b*=-.07, *t*(26.9)=-11.81, *p*<.001), such that CEN-SN density significantly decreased from baseline to follow-up. Within-CEN density did not change from baseline to follow-up in L/NA youth (*b*=-.002, *t*(26.5)=-.55, *p*=.58).

Of note, the post-hoc analyses that examined within-group change over time for the two density metrics with time by treatment interactions were reported with complete case data in the manuscript, in order to facilitate paired *t*-tests. However, these same post-hoc tests were also assessed in the larger sample using LMEs. These full usable sample LME models were run with covariates of time and mean FD. The results followed the same pattern of direction of change and significance, though the within-RMT model for CEN-SN did not converge. The results for those models are as follows: increase in CEN-SN density within EF-CBT, *b* = .03, *t*(66.7)=5.17, *p*<.001; decrease in CEN-SN density within RMT (did not converge); no change in within-CEN density with EF-CBT, *b*=-.003, *t*(68.3)=-.67, *p*=.51; increase in within-CEN density with RMT, *b*=.04, *t*(27.0)=8.32, *p*<.001.

**Supplementary Figures**

**Figure S1.** CONSORT Diagram for Usable Resting-state fMRI Data

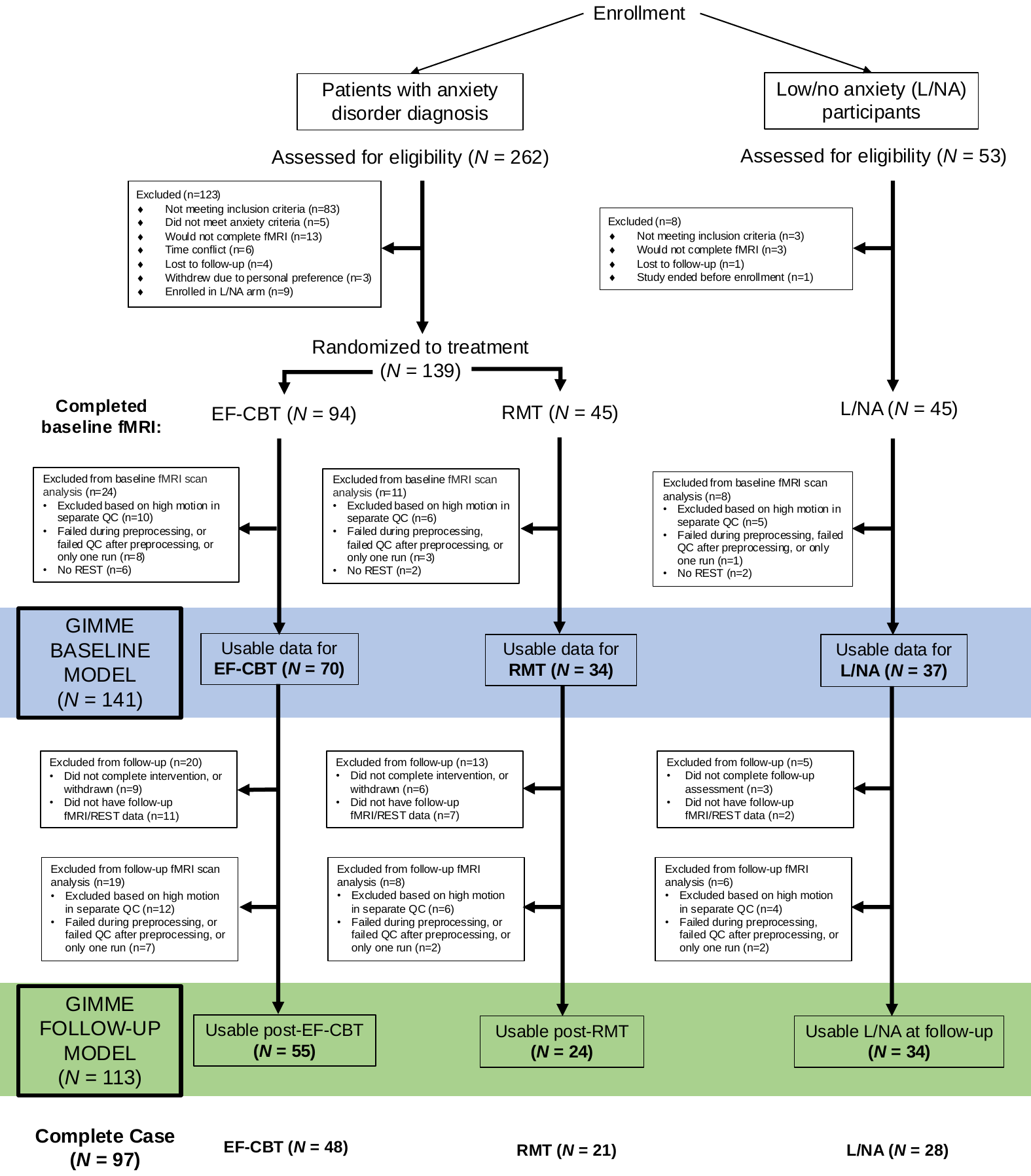

*Note.* CONSORT diagram demonstrates path of exclusions from the number of participants enrolled to those analyzed in the present analyses. EF-CBT=exposure-focused cognitive behavioral therapy; RMT=relaxation mentorship training; L/NA=low/no anxiety participants; rsfMRI=resting-state functional magnetic resonance imaging. The baseline GIMME model was run on participants highlighted in the blue bar; the follow-up GIMME model was run on those in the green bar. Complete case participants (bolded in the last row) had usable scans at baseline *and* follow-up (in the blue *and* green bars).

**Figure S2.** Example individual-level map from baseline of a participant assigned to EF-CBT

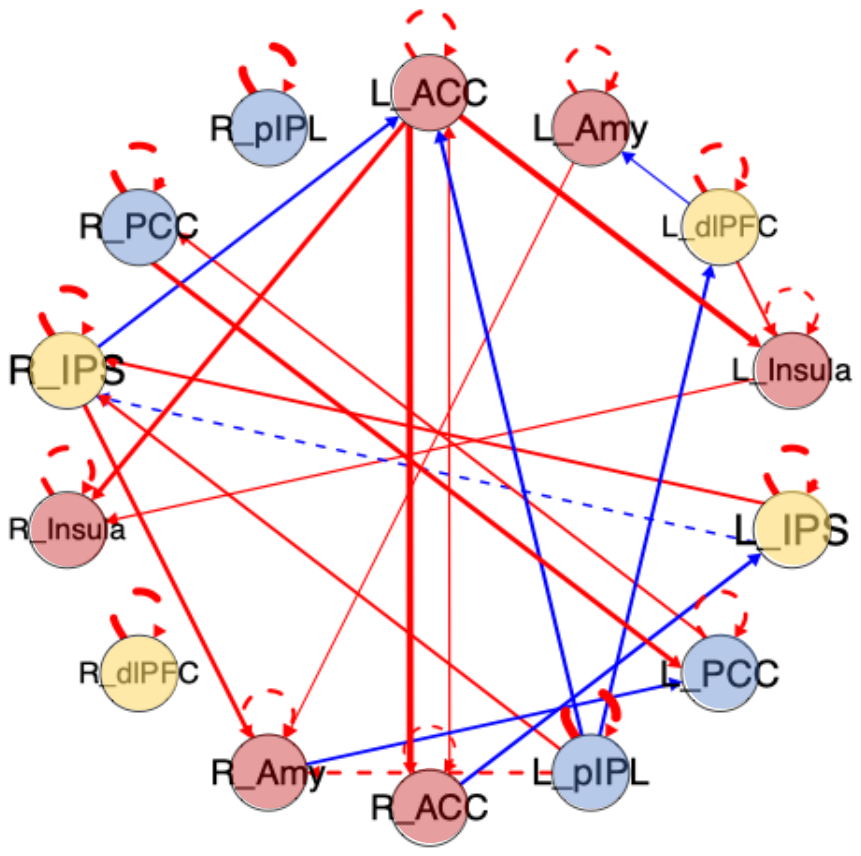

*Note.* Map generated by CS-GIMME to demonstrate connections at the individual-level for a single participant. Each circle represents a region-of-interest (ROI) and colors of each circle reflect *a priori* assigned network membership for each ROI: central executive network (CEN; yellow); salience network (SN; red); and default mode network (DMN; blue). Solid lines reflect contemporaneous connections, dashed lines reflect lagged connections, blue lines reflect negative connections, and red lines reflect positive connections. Line thickness reflects connection magnitude. R/L = right/left; ACC = anterior cingulate cortex; Amyg = amygdala; dlPFC = dorsolateral prefrontal cortex; Ins = insula; IPS = intraparietal sulcus; PCC = posterior cingulate cortex; pIPL = posterior inferior parietal lobule.

**Figure S3.** Change in Network Density by Treatment Group as Compared to L/NA Youth

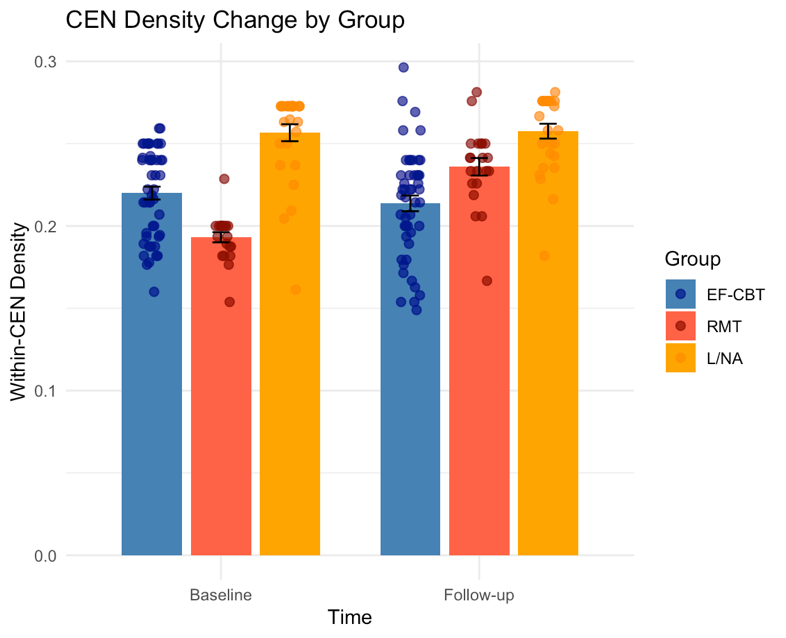

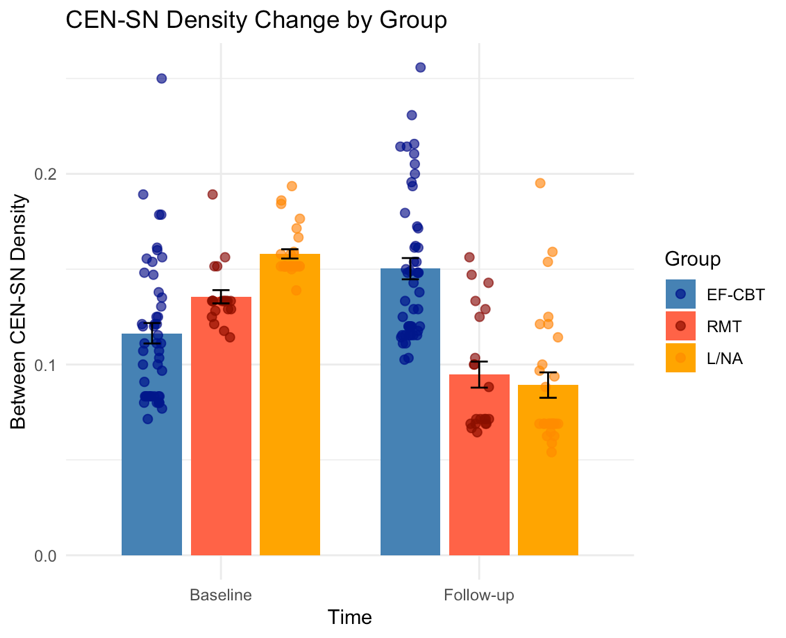

*Note.* Change in between-network CEN-SN and within-CEN density from baseline to follow-up, displayed by group (EF-CBT in blue, RMT in red, L/NA in orange). CEN*=*Central Executive Network; SN=Salience Network; EF-CBT=exposure-focused cognitive behavioral therapy; RMT=relaxation mentorship training; L/NA=low/no anxiety participants. Bars reflect the mean of each density metric and error bars indicate standard error. The dots on the bars are density values for each participant, color coded by the treatment to which they were assigned.

**Tables**

**Table S1.** Regions of Interest and Coordinates

| Brain Network | Region | x | y | z |
| --- | --- | --- | --- | --- |
| Central Executive | Right dorsolateral prefrontal cortex | 44 | 34 | 30 |
|  | Left dorsolateral prefrontal cortex | -44 | 34 | 30 |
|  | Right intraparietal sulcus | 34 | -46 | 42 |
|  | Left intraparietal sulcus | -34 | -46 | 42 |
| Default Mode |  |  |  |  |
|  | Right posterior inferior parietal lobule | 48 | -62 | 28 |
|  | Left posterior inferior parietal lobule | -48 | -64 | 28 |
|  | Right posterior cingulate cortex | 5 | -54 | 28 |
|  | Left posterior cingulate cortex | -5 | -52 | 28 |
| Salience |  |  |  |  |
|  | Right anterior cingulate cortex | 5 | 30 | 20 |
|  | Left anterior cingulate cortex | -5 | 30 | 20 |
|  | Right insula | 38 | 18 | -2 |
|  | Left insula | -36 | 18 | -2 |
|  | Right amygdala | 24 | -4 | -20 |
|  | Left amygdala | -24 | -4 | -20 |

**Table S2.** LME Models for Change in Density Over Time with L/NA Group as Reference

| **Outcome** |  | **Predictor** |  | ***Estimate*** | **SE** | ***df*** | **t** | ***p*** |
| --- | --- | --- | --- | --- | --- | --- | --- | --- |
| ***CEN-SN Density*** |  | Intercept |  | 0.17 | 0.01 | 213.17 | 14.69 | <.001 |
|  |  | Time |  | -0.07 | 0.01 | 134.14 | -10.27 | <.001 |
|  |  | EF-CBT |  | -0.05 | 0.01 | 246.00 | -7.32 | <.001 |
|  |  | RMT |  | -0.02 | 0.01 | 246.00 | -2.46 | 0.015 |
|  |  | Age |  | 0.00 | 0.00 | 172.27 | -2.13 | 0.035 |
|  |  | Framewise Displacement |  | 0.05 | 0.01 | 246.00 | 4.25 | <.001 |
|  |  | Time * EF-CBT |  | 0.10 | 0.01 | 134.76 | 11.94 | <.001 |
|  |  | Time * RMT |  | 0.03 | 0.01 | 139.42 | 2.64 | 0.009 |

| **Outcome** |  | **Predictor** |  | ***Estimate*** | **SE** | ***df*** | **t** | ***p*** |
| --- | --- | --- | --- | --- | --- | --- | --- | --- |
| ***Within-CEN Density*** |  | Intercept |  | 0.26 | 0.01 | 213.17 | 29.45 | <.001 |
|  |  | Time |  | 0.00 | 0.01 | 136.02 | -0.25 | 0.800 |
|  |  | EF-CBT |  | -0.04 | 0.00 | 246.00 | -8.09 | <.001 |
|  |  | RMT |  | -0.07 | 0.01 | 246.00 | -11.88 | <.001 |
|  |  | Age |  | 0.00 | 0.00 | 171.95 | 1.27 | 0.207 |
|  |  | Framewise Displacement |  | -0.07 | 0.01 | 246.00 | -8.29 | <.001 |
|  |  | Time * EF-CBT |  | 0.00 | 0.01 | 136.66 | -0.41 | 0.684 |
|  |  | Time * RMT |  | 0.04 | 0.01 | 141.41 | 5.30 | <.001 |

Results of models evaluating the interacting effect of treatment condition (EF-CBT, RMT; reference group is the L/NA group) and time (baseline, follow-up) on connectivity metrics of CEN-SN density and within-CEN density. Results are displayed for connectivity metrics where there was a significant treatment by time interaction (CEN-SN density and CEN density) in the primary analysis focused on comparing treatment groups (EF-CBT to RMT, directly). EF-CBT = Exposure focused Cognitive Behavioral Therapy; RMT = Relaxation Mentorship Training; CEN = Central Executive Network; SN = Salience Network.
